# Patient-centered care, advance care planning, and treatment preferences among home medical care patients in Japan: The ZEVIOUS study

**DOI:** 10.1101/2021.08.13.21261948

**Authors:** Shinu Hayashi, Yutaka Shirahige, Yukio Tsugihashi, Hidekazu Iida, Misaki Hirose, Masakazu Yasunaka, Noriaki Kurita, the ZEVIOUS group

## Abstract

**Background:** It remains unclear how both the quality of patient-centered care and the patient’s illness affect advance care planning (ACP) in primary care settings. Identifying the facilitators and barriers to ACP in primary care settings has become a growing scientific and clinical challenge.

**Objective:** To examine the association between the quality of primary care and ACP preparedness among patients. Additionally, to investigate whether ACP preparedness and the patient’s illness are associated with the expression of future treatment preferences.

**Design:** Multicenter cross-sectional study.

**Participants:** Adult Japanese patients receiving home medical care.

**Main Measures:** A survey was run to assess consideration of ACP by patients and expression of future treatment preferences. The quality of primary care, which reflects patient centeredness, was assessed with the Japanese version of the Primary Care Assessment Tool– Short Form. Information on the clinical conditions that required home medical care was collected from physicians.

**Key Results:** Of 194 patients using 29 home medical services, 62 patients (32%) showed signs of ACP preparedness, and 153 patients (78%) expressed their treatment preferences. In a multivariable-adjusted generalized estimating equation, primary care quality was associated with ACP preparedness (per 10-point increase, adjusted OR: 1.96, 95% CI: 1.51–2.56). However, we found insufficient evidence to support that ACP preparedness was associated with a lower incidence of non-expression of treatment preferences (adjusted OR: 1.02, 95% CI 0.49–2.12). In contrast, having cancer was associated with a lower incidence of non-expression of treatment preferences (adjusted OR: 0.12, 95% CI: 0.01–0.995).

**Conclusions:** At a minimum, patient centeredness in home medical care facilitates must ensure the initiation of ACP preparedness. To understand the association between ACP preparedness and expression of treatment preferences, further efforts are warranted to clarify the quality and content of ACP preparedness simultaneously with the patient’s illness.

## Introduction

Advance care planning (ACP) is the process whereby the patient, their families, and healthcare professionals discuss and prepare for future treatments that are consistent with the patient’s personal values, goals, and preferences in case a treatment decision is needed, including at the end of life [1]. ACP is known to have positive effects on the process and outcomes related to end-of-life care, including reduced invasive use of cardiopulmonary support measures, improved quality of life for the patient and their family [2], increased satisfaction of the patient and their family [3], and reduced stress, anxiety, and depression for the bereaved family [4]. In the primary care setting, identifying promoters and barriers to ACP have become urgent challenges [5]. To date, consensus recommendations include interactive and repeated discussions between the patient and their physician about ACP that reflect the patient’s life stage [5]. The quality of primary care is a potential area to target for promoting ACP, given the emphasis on patient-centered care in the assessment of primary care quality and the fact that both the process and quality of ACP are based on respecting the patient’s treatment preferences and values. [6,7] Despite the theoretical importance of primary care quality, evidence on the potential benefit that the quality of primary care may bring to ACP preparedness is very limited.

A previous study found that the quality of primary care assessed through the patient experience was associated with ACP discussions in an outpatient primary care setting [8]. However, ACP preparedness is less urgent in such outpatient settings. Most of the study participants were aged under 80, rated their health as good at least, and did not experience ACP discussions [8]. ACP preparedness is a more urgent issue for patients who are unable to visit a clinic because of advanced age, disability, or life-threatening morbidities. In Japan, those patients typically receive home medical care with regular visits from their physicians [9]. Thus, clarifying the influence of the quality of the primary care provided by these physicians on their patients’ ACP preparedness, and the impact of this preparedness on patients’ treatment preferences regarding life prolongation, will provide important insights for a superaged country like Japan, with a large population likely to require home medical care in the future.

Therefore, we conducted a multicenter cross-sectional study using data from the Zaitaku Evaluative Initiatives and Outcome Study (ZEVIOUS) to examine the implementation of ACP preparedness and its association with the quality of primary care among patients receiving home medical care. In addition, we investigated potential barriers to ACP and the association between ACP preparedness and the expression of patients’ values around life prolongation.

## METHODS

### Design, setting, and participants

ZEVIOUS was a multicenter cross-sectional study involving 29 home medical care facilities located in the Tokyo Metropolitan area, Nara, and the Nagasaki Prefecture in Japan. Patients were eligible if they (1) were receiving home medical care by home-care physicians working at one of the participating facilities; and (2) were judged by their physicians as being capable of responding to the questionnaire survey. The questionnaire was administered to each patient between January and July 2020 and the patients were asked to complete it at home. Patients who could not write because of visual impairment or physical disability were allowed to complete the form with the aid of a family member or a formal caregiver. In the questionnaire, we provided the instruction that all questions should be answered based on the patient’s personal views, and we gave assurances that the responses would not be reviewed by their treating physicians. The completed questionnaires were sent to a central research institute for analysis. This study was approved by the institutional review board of Fukushima Medical University (ippan-30254).

### Conceptual framework

A conceptual framework for our study is shown in Figure S1. First, we focused on the relationship between the patient experience, which was considered both as an exposure and an indicator of the quality of primary care, and preparedness for ACP as an outcome (analysis #1). Second, we focused on the relationship between ACP preparedness as an exposure and the non-expression of treatment preferences on life prolongation as an outcome (analysis #2).

### Patient experience as an indicator of primary care quality

The Japanese version of Primary Care Assessment Tool–Short Form (JPCAT–SF) was used to measure the patient experience among patients receiving home medical care. [10] This instrument is a short version of the original 29-item JPCAT [11], which was itself an adaptation to the Japanese culture of the Primary Care Assessment Tool (PCAT) designed to measure the experience of adult patients in primary care. [12] The JPCAT–SF is a 13-item tool comprising six domains representing five primary care attributes, including first contact (two items), longitudinality (two items), coordination (three items), comprehensiveness (two items for the services available and two items for the services provided) and community orientation (two items) [10]. Details of the items and domains of the JPCAT-SF are presented in Supplementary Item S1 and Item S2. For each item, participants were asked to respond on a 5-point Likert scale, ranging from *strongly disagree* to *strongly agree*. Each response was converted to an item score ranging from 0 to 4. The domain scores were calculated by multiplying the average of the item scores in the same domain by 25 (i.e., ranging from 0 to 100), with higher scores indicating better performance. In the coordination domain, which asks about experiences with referrals to a specialist, respondents who had never seen a specialist were given 50 points (the midpoint of all the possible scores). The total score was the average of the six domain scores and represented an overall measure of the patient experience of primary care. The JPCAT–SF has been shown to have good internal-consistency reliability (Cronbach’s α = 0.77 for the total score, Cronbach’s α > 0.76 for each domain score) and excellent criterion validity (Pearson correlation coefficient with the original 29-item JPCAT and the overall rating for usual care facilities: 0.94 and 0.43, respectively) [10].

### Preparedness for advance care planning

In line with a treatment preference questionnaire administered for end-stage renal disease patients in the United States [13], the following instruction statement was provided: “This section asks about thoughts on your health care if you were to become very sick in the future.” Then, the following question was asked: “Have you thought about the kinds of treatments you would want or not want if you were to become very sick and were unable to speak in the futureã (Check all items that apply).” Multiple choices were provided, consisting in the following responses: “I have not thought about this,” “I have thought about this, but have not talked about it to a family member, others (including a friend), or my doctor,” “I have talked about this with a family member or others (including a friend),” “I have talked about this with my doctor,” or “I have written a document or memo about my preferences.” We defined the presence of ACP preparedness when respondents chose either “I have talked about this with my doctor” or “I have written a document or memo about my preferences,” because recent studies on ACP interventions have employed a combination of communication and establishment of advance directives [14,8].

### Reasons not to communicate about advance care planning

Respondents who chose “I have thought about this, but have not talked about it to a family member, others (including a friend), or my doctor” were asked to answer the next question: “Please tell us why. (Check all items that apply.)” Multiple choices were provided, consisting in the following responses: “I don’t want to talk about it,” “I don’t feel the need to talk about it,” “I didn’t have a chance to talk about it,” “I don’t know what to discuss because of lack of knowledge,” “I don’t have anyone to talk to,” or “Other.”

### Choices about treatment preferences

In line with the aforementioned questionnaire on treatment preferences [13], the following question was asked: “If you were to become very sick in the future and were unable to speak, would you prefer a medical care that focuses on extending life as much as possible (even though it may increase pain and distress), or would you want a medical care that focuses on relieving pain and distress (even though it may not extend life)ã (Check only one answer.)” A single choice was allowed among the following responses: “I prefer medical care that extends life (even if it may increase pain and distress),” “I prefer medical care that relieves pain and distress as much as possible (even if it does not prolong life),” or “I am not sure which I would choose.” Since the goal of ACP is to establish treatment preferences based on the patient’s own values, we defined a negative outcome as no decision made about treatment preferences (i.e., choosing “I am not sure which I would choose.”).

### Covariates

Information about age, sex, education, and presence of a family member were collected through the questionnaire. Data on the presence of comorbidities (primary and others) for which home medical care was required, the type of residence, and the patient’s life expectancy (less than 1 year or not) were provided by the treating physicians.

### Statistical analysis

Statistical analyses were conducted using Stata/SE, version 15 (Stata Corp., College Station, TX, U.S.A.). Patient characteristics were described as means and standard deviations for continuous variables and numbers and proportions for categorical variables. The numbers and proportions of patients’ responses regarding preparedness for ACP were analyzed. Among those who answered they had never talked to their family or doctor about ACP, the numbers and proportions of responses regarding the reasons for this were analyzed.

To estimate the association between JPCAT–SF scores and the likelihood of ACP preparedness, we fit a series of generalized estimating equations under an exchangeable working correlation structure (analysis #1). The rationale for choosing this model was to estimate odds ratios while addressing facility-level clustering effects [15]. Covariates included age, gender, education, presence of family, prognostic expectation, dementia, cancer, and weakness. In addition to the model in which the total JPCAT–SF score was treated as an explanatory variable, we also fit models in which each of the six domain scores of JPCAT–SF was treated as an explanatory variable. To estimate the predicted probabilities for ACP preparedness across the JPCAT–SF total score, we calculated the probabilities standardized to the total study population with all other variables set to their original values [16]. Similarly, to estimate the association between ACP preparedness and the likelihood of not expressing a treatment preference, we fit a series of generalized estimating equations under an exchangeable working correlation structure (analysis #2). Within each analysis, missing data on covariates, which were assumed to be missing at random, were addressed through five imputations using a chained equations method [17]. P < 0.05 was considered to indicate statistical significance.

## RESULTS

### Patient characteristics

Of the 202 patients who received home medical care, 8 patients who did not have a JPCAT– SF score were excluded, leaving a total of 194 patients for the analyses. The patients’ characteristics are presented in Table 1. The mean age (standard deviation) was 80.0 (14.1) years; 116 patients (60%) were women; and 176 patients (91%) lived at home. Regarding the comorbidities requiring home medical care, 26 patients (13%) had cancer, 37 patients (19%) had dementia, and 36 patients (19%) had weakness associated with advanced age. Thirty-five patients (18%) were expected to live less than one year.

**Table 1.**
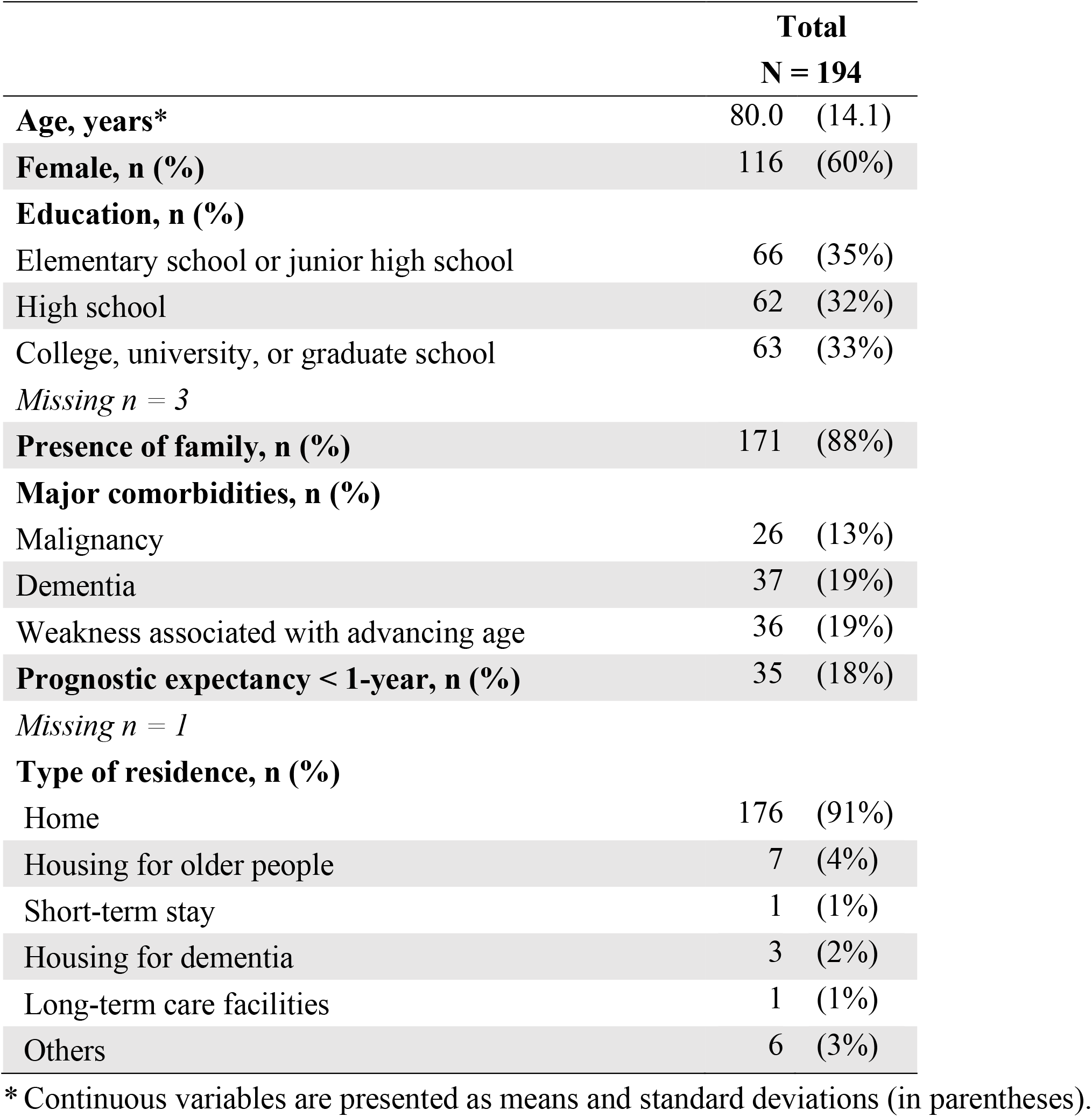
Characteristics of the patients with home care medicine.

### Preparedness for ACP and reasons for not discussing ACP

Patients’ responses about preparedness for ACP are shown in Supplementary Table S1. Discussion with the family physician was reported in 57 patients (29%), and preparation of a written document was reported by 21 patients (11%). The total number of respondents who had been involved in ACP activities was 62 (32%). On the other hand, there were 76 patients (39%) who had never thought about the treatment they would like to receive or not receive, and 50 patients (26%) who had thought about treatment but had never discussed it with family or family physicians.

The responses to the reasons for not discussing ACP with family members or physicians are shown in Table 2. A lack of opportunities was the most common reason reported by 24 patients (48%). Eighteen patients (36%) did not feel the need to discuss ACP, and 13 patients (26%) did not know what to discuss due to a lack of knowledge. Three patients (6%) answered that they did not want to talk about it.

**Table 2.** Reasons for not having talked to family members or their doctors (n = 50)

| **Reason, n (%)\*** |  |  |
| --- | --- | --- |
| I didn't have a chance to talk about it. | 24 | (48%) |
| I don't feel the need to talk about it. | 18 | (36%) |
| I don't know what to discuss because I don't have knowledge. | 13 | (26%) |
| I don't want to talk about it. | 3 | (6%) |
| I don't have anyone to talk to. | 3 | (6%) |
| Others | 4 | (8%) |
| I don't understand this disease very well. | 1 |  |
| I don't know whether the treatments I would like to receive are right for my condition. | 1 |  |
| I will follow my family's opinion. | 1 |  |

\* Multiple choices are allowed.

### Quality of primary care services and its relationship with preparedness for ACP

The mean JPCAT–SF total score was 70.9 (15.6), with the lowest and highest scores of 25 and 100, respectively. The association between the JPCAT–SF total score and preparedness for ACP is shown in Table 3. A higher JPCAT–SF total score was associated with a higher likelihood of preparedness for ACP (adjusted OR for every 10-point increase: 1.96, 95% CI: 1.51–2.56). There was insufficient evidence to conclude that cancer (adjusted OR: 2.31, 95% CI: 0.80–6.68) and dementia (adjusted OR: 0.76, 95% CI: 0.31–1.83) were associated with a higher likelihood of preparedness for ACP.

**Table 3.**
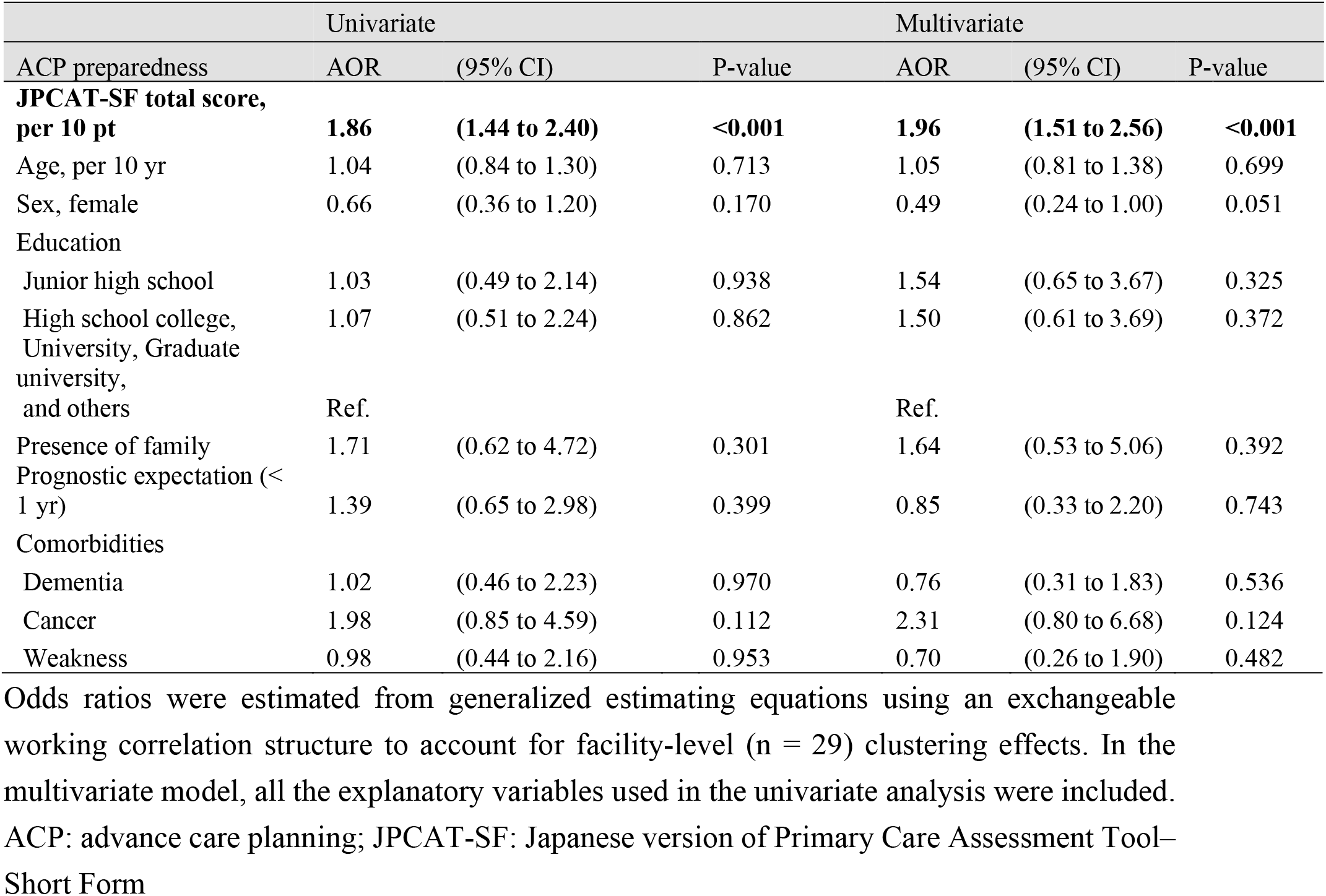
Factors associated with preparedness for advance care planning (n = 194)

The probabilities of preparedness for ACP standardized to the total study population across JPCAT–SF total scores is shown in Figure 1. The probabilities of preparedness for ACP increased as the JPCAT–SF total score increased. At the JPCAT–SF total scores of 40, 70, and 100 points, the probabilities of preparedness for ACP were 5.0% (95% CI: 0.5%–9.6%), 27.3% (95% CI: 20.7%– 34.0%), and 71.8% (95% CI: 58.0%–85.6%), respectively.

**Figure 1.**
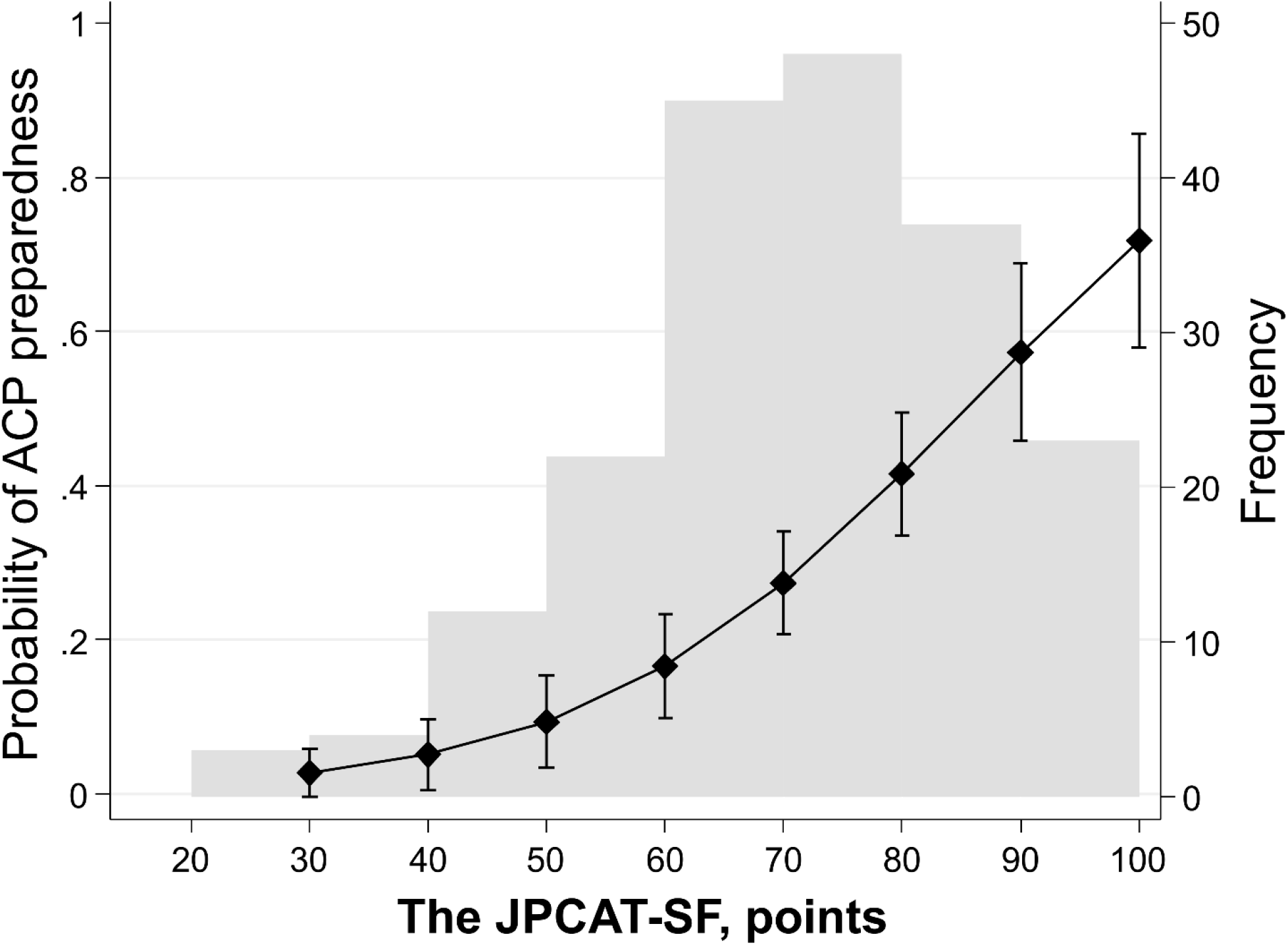
Probability of preparedness for advanced care planning estimated by the JPCAT–SF total score. Using the multivariable generalized estimating equation model, adjusted probability by the JPCAT–SF total score was predicted. The left vertical axis shows probability of preparedness for ACP. The solid circle indicates point estimates. The vertical lines indicate 95% confidence intervals. Gray bars indicate frequency of the JPCAT–SF total score values. The right vertical axis shows frequency of each gray bar. n = 194. ACP: advance care planning; JPCAT–SF: The Japanese version of Primary Care Assessment Tool–Short Form.

The association between each domain of the JPCAT–SF questionnaire and preparedness for ACP is shown in Supplementary Table 2. In all cases where first contact, longitudinality, coordination, services available, services provided, and community orientation were used as an individual exposure variable, a higher score was associated with a higher likelihood of preparedness for ACP. The adjusted OR (95% CI) per 10-point increase in each domain were 1.36 (1.13–1.64), 1.32 (1.07–1.62), 1.19 (1.05–1.35), 1.22 (1.03–1.43), 1.19 (1.09–1.31), and 1.34 (1.14–1.57), respectively.

### Association between preparedness for ACP and treatment preference

Patients’ responses about their treatment preferences in the case of having difficulty in expressing their willingness in future are shown in Supplementary Table 3. There were 141 patients (75%) who preferred palliating symptoms, 7 patients (4%) who preferred prolonging life, and 41 patients (22%) who were unable to decide their preference.

The relationship between being unable to decide their preference as an outcome and patients’ preparedness for ACP is shown in Table 4. There was insufficient evidence to demonstrate that preparedness for ACP was associated with the expression of treatment preferences (adjusted OR: 1.02, 95% CI: 0.49–2.12). On the other hand, patients with cancer were significantly more likely to express a preference (adjusted OR: 0.12, 95% CI: 0.01–0.995).

**Table 4.** Factors associated with non-expression of treatment preferences (n = 189) Odds ratios were estimated from generalized estimating equations using an exchangeable

| Symptom management | Univariate |  |  | Multivariate |  |  |
| --- | --- | --- | --- | --- | --- | --- |
|  | AOR | (95%CI) | P-value | AOR | (95%CI) | P-value |
| ACP preparation, present | 0.97 | (0.46 to 2.02) | 0.925 | 1.02 | (0.49 to 2.12) | 0.949 |
| Age, per 10 yr | 1.02 | (0.80 to 1.29) | 0.900 | 0.98 | (0.76 to 1.26) | 0.843 |
| Sex, female | 0.61 | (0.32 to 1.14) | 0.123 | 0.62 | (0.30 to 1.28) | 0.197 |
| Education |  |  |  |  |  |  |
| Junior high school | 1.40 | (0.64 to 3.07) | 0.405 | 1.40 | (0.61 to 3.24) | 0.425 |
| High school | 0.47 | (0.19 to 1.20) | 0.116 | 0.58 | (0.23 to 1.50) | 0.261 |
| College, University, Graduate university, and others | Ref. |  |  | Ref. |  |  |
| Presence of family | 1.62 | (0.53 to 4.95) | 0.398 | 1.84 | (0.58 to 5.87) | 0.304 |
| Prognostic expectation (< 1yr) | 0.61 | (0.23 to 1.61) | 0.321 | 0.93 | (0.33 to 2.62) | 0.896 |
| Comorbidities |  |  |  |  |  |  |
| Dementia | 1.10 | (0.48 to 2.55) | 0.820 | 1.13 | (0.45 to 2.86) | 0.794 |
| **Cancer** | **0.11** | **(0.01 to 0.84)** | **0.034** | **0.12** | **(0.01 to 0.995)** | **0.050** |
| Weakness | 1.38 | (0.61 to 3.12) | 0.443 | 1.20 | (0.47 to 3.06) | 0.705 |

working correlation structure to account for facility-level (n = 29) clustering effects. In the multivariate model, all the explanatory variables used in the univariate analysis were included.
ACP: advance care planning

## Discussion

This study showed that the quality of primary care experience was associated with ACP preparedness among patients receiving home medical care. However, we found insufficient evidence to demonstrate that ACP preparedness was associated with patients’ expression of treatment preferences. We hope that these findings will serve as a guide for the successful initiation and continuation of ACP in home medical care settings.

Our study findings are consistent with and extend previous research findings in outpatient primary care settings on the association between the quality of primary care measured as patient experience and preparedness for ACP [8]. First, about 30% of the patients in our study had discussed ACP with their physicians, which was much higher than the proportion in the outpatient primary care setting study [8]. Specifically, two-thirds of the outpatient primary care patients were in their 60s and 70s, and more than 80% rated their health as good and thus seemed to be less likely to discuss ACP. In contrast, the patients in our study were 80 years old on average and included patients with limited life expectancy or functioning to the extent that they could not visit a clinic by themselves. Therefore, our findings are more applicable to home medical care patients faced with the possible need to prepare ACP than the findings from previous studies.

Second, more than 20% of the patients in our study had made no decision about their future treatment preferences, well above than the 3.2% of residents in Southern California nursing homes who expressed no preference [18]. The possible reasons for this difference include the cultural values of family-centered decision-making in Asia versus the values of individual autonomy in Europe and the United States [19,20]. A passive attitude toward ACP discussions and natural preference for physician-led ACP has also been reported, especially in Japanese people [21,22]. In addition, the lack of a legal framework for ACP consisting, for example, in a mandate to offer patients autonomy regarding their living will and exemption to physicians for implementing decisions specified in advance directives, may be another possible factor.

Third, the target population for examining ACP preparedness in this study included a variety of diseases that led to the initiation of home medical care. This allowed us to confirm that having cancer is associated with a greater expression of future treatment preferences compared to not having cancer. Unsurprisingly, this finding may be due to an inevitable consideration of death through personal experience with cancer care.

Our study has several implications for physicians and researchers alike. First, the associations between each domain of patient experience and ACP preparedness indicate that a greater commitment to patient centeredness in home medical care may provide more opportunities to initiate ACP preparedness. For example, experience of out-of-hours medical care for new needs, measured as first contact, can lead to greater confidence between the patient and their physician and prompt a discussion about future treatment plans in case of unexpected health deterioration. The holistic consideration of a patient, measured as longitudinality, can serve as a basis for smoother communication about ACP preparedness. For example, a physician who knows a patient’s experience of major surgery can reflect together with them and discuss future invasive procedures in detail [23]. A doctor who knows about family dynamics can encourage the patient to think about ACP in order to maintain a good family relationship [24]. The appropriate referral to a specialist (e.g., oncologist or geriatrician), measured as coordination, will allow both a patient and their physician to correctly understand their medical condition and their clinical prospects, providing a basis for a specific and realistic future treatment plan [25,1]. Advice on health literacy and supplements, measured as the comprehensiveness of the services provided, can facilitate ACP preparedness, as it allows a patient and their physician to discuss appropriate modalities for symptom management, including at the end of life. Considering the needs of community patients, measured as community orientation, as well as the effective use of constrained local healthcare resources, a physician may bring in a timely discussion of pragmatic options for end-of-life care. Whether the development of educational and training programs for home medical care that reflects these patient-centeredness domains will improve the quality of home medical care warrants further investigation.

Second, the insufficient evidence supporting that ACP preparedness was associated with the expression of treatment preferences, while ACP preparedness was found to be associated with cancer, should encourage physicians and researchers to deliberate more on the reasons for non-expression. One possible reason why ACP discussions do not necessarily lead to expression of treatment preferences is that the physicians who engage in the discussion may be trying to find the right time for a patient to specify their treatment preferences based on their health status [1]. For example, a patient without a life-threatening illness may feel that the ACP discussion was not the right time to express a preference [26] and may choose not to decide their preference as an active strategy to cope with an uncertain situation [27], which their physician may accept as an evolving process.

Furthermore, the ACP discussion will need to demonstrate quality and depth in order to encourage patients to express treatment preferences. In addition to the communication skills embedded in the principles of patient centeredness, video decision aids and dialog support tools need to be developed [28,29]. On the other hand, the reason why cancer is associated with expression of treatment preferences may be attributable to patients’ experiences of choosing their treatment preferences during cancer care, regardless of discussions with home medical care physicians.

This study has several strengths. First, this is the first study to demonstrate that the quality of the primary care experience in home medical care, as measured with a psychometrically-validated scale, is associated with ACP preparedness upon accounting for facility clustering effects. Second, since our study was conducted in a multi-center home medical care setting in rural and urban areas in Tokyo, Nara, and Nagasaki, our findings are relatively generalizable to other settings and populations.

This study also has several limitations. First, the largely positive patient experience may not be representative of the national level of home medical care, as this project was initiated by a group of physicians who wish to improve the quality of home medical care. Second, the primary care quality and ACP preparedness among patients who were considered unable to answer the questionnaire remain unknown. Third, as in other studies, no data on depth or quality of ACP preparedness was collected [30]. Fourth, the relationship between the quality of patient experience and ACP preparedness observed in this study may appear to be self-evident, since ACP is one of the items contained in the JPCAT–SF. However, this potential criticism was addressed by simultaneously demonstrating the relationship between each domain of the JPCA–SF and ACP preparedness. Alternatively, it could be argued that we were able to demonstrate the criterion validity of the qualitative scale in home medical care settings by showing the relationship between the total JPCAT–SF score and ACP preparedness [31].

In conclusion, ACP preparedness was reported by about 30% of the home medical care patients who were able to respond to the questionnaire. Furthermore, a higher quality of primary care experience was associated with a higher likelihood of ACP preparedness. These findings indicate that patient-centered care in home medical care settings can at least encourage the initiation of ACP preparedness. Further research is warranted to determine whether the quality and depth of physicians’ activities towards ACP preparedness contribute to patients’ expression of treatment preferences.

## Data Availability

The datasets used and/or analyzed during the current study are available from the corresponding author on reasonable request.

## Contributors

The authors would like to express their sincere gratitude to the research assistants Ms Miyuki Sato and Ms Lisa Shimokawa (Fukushima Medical University Hospital, Fukushima-city, Fukushima) for their assistance in collecting the questionnaire-based information used in this study.

