## Supplemental for "Patient-centered care, advance care planning, and treatment preferences among home medical care patients in Japan: The ZEVIOUS study"

**Appendices**

**Figure S1. Study conceptual framework**
ACP: Advance care planning; JPCAT-SF: The Japanese version of Primary Care Assessment Tool–Short Form

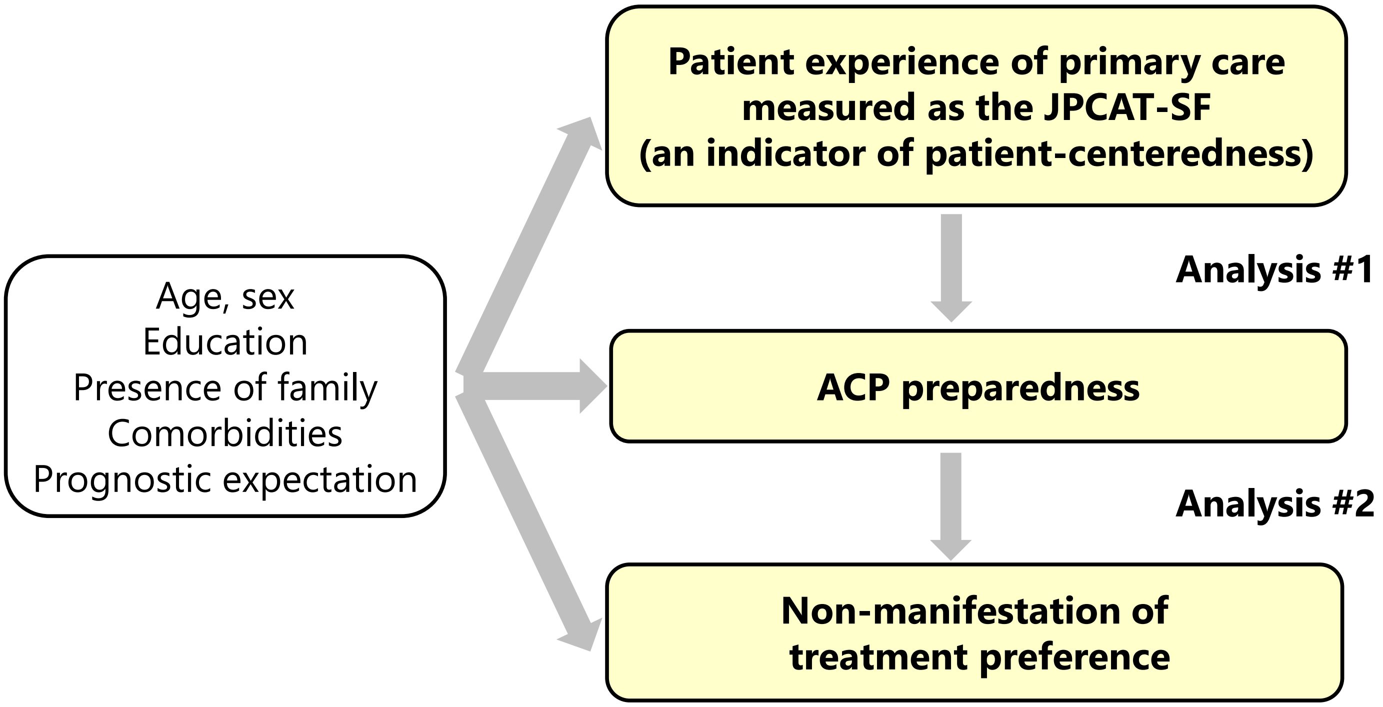

**Supplementary Instruction S1. Items and responses for the JPCAT-SF [1]**

Questionnaires in Japanese version are available from the following website(https://bfffe681-45f7-48c3-aa93-2f67612434a5.filesusr.com/ugd/6c0e9c_7c80f3e3ce6e45eebb5a3aedfb9a2800.pdf)

| | |
|---|---|
| Instruction sentences | (English: “Check the box that best fits each question.”) |
| Question 1 | (English: “When your Primary Care Practice is closed on Saturday and Sunday and you get sick, would someone from there see you the same day?”) |
| Response to Question 1 | (English: “Strongly agree/Somewhat agree/Not sure/Somewhat disagree/Strongly disagree ”) |
| Question 2 | (English: “When your Primary Care Practice is closed and you get sick during the night, would someone from there see you that night?”) |
| Response to Question 2 | (English: “Strongly agree/Somewhat agree/Not sure/Somewhat disagree/Strongly disagree ”) |
| Question 3 | (English: “Does your Primary Care Physician (PCP) know you very well as a person, rather than as someone with a medical problem?”) |
| Response to Question 3 | (English: “Strongly agree/Somewhat agree/Not sure/Somewhat disagree/Strongly disagree ”) |
| Question 4 | こ(English: “Does your PCP know what problems are most important to you?”) |
| Response to Question 4 | (English: “Strongly agree/Somewhat agree/Not sure/Somewhat disagree/Strongly disagree ”) |
| Question 5 | (English: “Have you ever had a visit to a specialist or special service of any kind?”) |
| Response to Question 5 | (English: “Yes/No or Not sure”) |
| Question 6 | (English: “Did your PCP suggest you go to the specialist or special service?”) |
| Response to Question 6 | (English: “Strongly agree/Somewhat agree/Not sure/Somewhat disagree/Strongly disagree ”) |
| Question 7 | (English: “Did your PCP discuss with you the different places you could have visited to get help with that problem?”) |
| Response to Question 7 | (English: “Strongly agree/Somewhat agree/Not sure/Somewhat disagree/Strongly disagree ”) |
| Question 8 | (English: “Please indicate whether it is available at your PCP’s office. Counseling related to abuse”) |
| Response to Question 8 | (English: “Strongly agree/Somewhat agree/Not sure/Somewhat disagree/Strongly disagree ”) |
| Question 9 | (English: “Please indicate whether it is available at your PCP’s office. Counseling related to personal preferences about end-of-life issues”) |
| Response to Question 9 | (English: “Strongly agree/Somewhat agree/Not sure/Somewhat disagree/Strongly disagree ”) |
| Question 10 | (English: “In visits to your PCP, are any of the following subjects discussed with you? Advice about over-the-counter medications.”) |

| | |
|---|---|
| Response to Question 10 | (English: “Strongly agree/Somewhat agree/Not sure/Somewhat disagree/Strongly disagree ”) |
| Question 11 | (English: “In visits to your PCP, are any of the following subjects discussed with you? Advice about medical information in the media: on TV, in the newspaper, etc.”) |
| Response to Question 11 | (English: “Strongly agree/Somewhat agree/Not sure/Somewhat disagree/Strongly disagree ”) |
| Question 12 | (English: “Does your PCP investigate whether the available health care is meeting the needs of the community?”) |
| Response to Question 12 | (English: “Strongly agree/Somewhat agree/Not sure/Somewhat disagree/Strongly disagree ”) |
| Question 13 | (English: “Does your PCP investigate the concerns people have about health problems in your community?”) |
| Response to Question 13 | (English: “Strongly agree/Somewhat agree/Not sure/Somewhat disagree/Strongly disagree ”) |

Each domain consists of the following items:
First contact domain - Questions 1 and 2
Longitudinality domain - Questions 3 and 4
Coordination domain - Questions 5, 6, and 7
Comprehensiveness (services available) domain - Questions 8 and 9
Comprehensiveness (services provided) domain - Questions 10 and 11
Community orientation domain - Questions 12 and 13

**Supplementary Item S2. Description of concepts of the JPCAT-SF domains**

First contact
Care is first sought from a primary care provider when a new health or medical need arises. The service should also be accessible and usable by the population as a new need or problem arises [1]. First contact is closely related to "access to care," a domain of patient-centered care characterized by the timely availability of care that is tailored to the patient [2]. The JPCAT–SF mainly measures patient experience related to off-hours care in primary care [1].

Longitudinality
It refers to the longitudinal use of usual sources of care, regardless of illness or injury [1]. Longitudinality is supported by one of the principles for patient centeredness, namely the consideration of the "patient as a unique person," i.e., the primary care physician's recognition of the patient's uniqueness (individual needs, preferences, values, beliefs, concerns, etc.) [2]. The JPCAT–SF mainly measures whether a patient feels that their primary care physician recognizes them as a whole person [1].

Coordination
The essence of coordination is the availability of information about past and existing problems and services, and the recognition of that information in relation to a current care need [1]. It relates to "coordination and continuity of care," which is an enabler of patient-centered care, i.e., facilitation of care that is well coordinated and continuous [2]. The JPCAT–SF mainly measures patient experience regarding past referral to a specialist [1].

Comprehensiveness (services available)
It refers to the availability of a wide range of services by a primary care provider and their appropriateness for a spectrum of needs for all but the most uncommon problems [1].
Under "services available," the JPCAT–SF mainly measures whether a patient feels they can receive care for mental health, dementia, and advanced care planning, if necessary [1].

Comprehensiveness (services provided)
It includes appropriate advice on daily lifestyle habits, self-medication, and health literacy [1,3]. It is underpinned by patient empowerment, an activity of patient centeredness, in which a primary care physician recognizes and actively supports a patient's ability and responsibility to self-manage their illness. [2] The JPCAT–SF mainly measures patient experience in terms of whether they received such appropriate advice.

Community orientation
It refers to care that is delivered in the context of the community [1] and is considered as a derivative domain of principles of primary care [4].
The JPCAT–SF mainly measures patient experience regarding home visits and whether a patient feels that their primary care physician is interested not only in their individual health problem, but also in problems in the community [1].

**Supplementary Table S1. Preparedness for advance care planning**

| | | |
|---|---|---|
| **Preparedness, n (%)*** | | |
| I have talked with family members or others (including friends) about the medical care I would like to receive or not receive. | 90 | (46%) |
| I haven't thought about the medical care I would like to receive or not receive. | 76 | (39%) |
| I have talked with my doctor about the medical care I would like to receive or not receive. | 57 | (29%) |
| I've thought about the medical care I would like to receive or not receive, but I haven't discussed it with family members or others (including friends) or my doctor | 50 | (26%) |
| I have written documents or notes about the medical care I want to receive or not receive. | 21 | (11%) |
| **Having talked with my doctor or written documents or notes, n (%)** | | |
| Yes | 62 | (32%) |

n = 194. * Multiple choices are allowed.

**Supplementary Table S2. Associations between domains of patient experience and advance care planning preparedness (n = 194)**

| | Univariate | | | Multivariate | | |
|---|---|---|---|---|---|---|
| ACP preparedness | AOR | (95% CI) | P-value | AOR | (95%CI) | P-value |
| **Domains of Patient Experience** | | | | | | |
| First contact, per 10 pt | **1.36** | **(1.13 to 1.64)** | **0.001** | **1.38** | **(1.14 to 1.67)** | **0.001** |
| Longitudinality, per 10 pt | **1.32** | **(1.07 to 1.62)** | **0.009** | **1.34** | **(1.08 to 1.65)** | **0.008** |
| Coordination, per 10 pt | **1.19** | **(1.05 to 1.35)** | **0.006** | **1.19** | **(1.05 to 1.35)** | **0.007** |
| Comprehensiveness (services available), per 10 pt | **1.22** | **(1.03 to 1.43)** | **0.018** | **1.22** | **(1.03 to 1.44)** | **0.022** |
| Comprehensiveness (services provided), per 10 pt | **1.19** | **(1.09 to 1.31)** | **<0.001** | **1.20** | **(1.09 to 1.32)** | **<0.001** |
| Community orientation, per 10 pt | **1.34** | **(1.14 to 1.57)** | **<0.001** | **1.36** | **(1.15 to 1.60)** | **<0.001** |

Odds ratios were estimated from generalized estimating equations using an exchangeable working correlation structure to account for facility-level (n = 29) clustering effects. In the multivariate model, the explanatory variables used in the univariate analysis and covariates (i.e., age, gender, education, presence of family, prognostic expectation, dementia, cancer, and weakness) were included.
ACP: advance care planning

**Supplementary Table S3. Treatment preferences in future when you are unable to speak due to a severe illness**

| | | |
|---|---|---|
| Prefer medical care that prolongs life (even if it may increase pain and distress) | 7 | (4%) |
| Prefer medical care that relieves as much pain and distress as possible (even if it doesn't extend life) | 141 | (75%) |
| I don't know which one to choose | 41 | (22%) |
| *Missing n = 5* | 1 | |

**Supplementary Text S1. Full details of the other members of the ZEVIOUS Group**

Shinsuke Muto, MD, PhD, EMBA, MPH[1]; Tatsunobu Natsubori, MD, PhD[1,2]; Michiko Hinata, MD[1,3]; Wataru Nakagawa, MD[1]; Akihiko Yonenaga, MD[1]; Lina Inagaki, MD, PhD[1]; Shioto Itakura, MD[1]; Nobuhiro Ikeda, MD, PhD[1]; Tomoka Nakamura, MD, PhD[1]; Naoya Miyashita, MD[1]; Takuya Furugen, MD[1]; Takafumi Abo, MD, PhD[4,5]; Sadayuki Okudaira, MD, PhD[4,6]; Kazuhiko Takuma, MD[4,7]; Chihiro Tsuchiya, MD, PhD[4,8]; Masahiro Deguchi, MD[4,9]; Takashi Fujii, MD, PhD[4,10]; Yoshitaka Harada, MD, PhD[4,11]; Seiji Matsuo, MD[4,12]; Motomichi Nakagawa, MD, PhD[4,13]; Ken Tanigawa, MD, PhD[4,14]; Yoshio Ochi, MD[4,15]; Sadanobu Ogasawara, MD[4,16]; Kazuhiko Hoshino, MD, PhD[4,17]; Momoko Aruga, MD[18]; Yoshinori Nakamura, MD[18]; Nobuhiro Sawa, MD[19]; Yosuke Akashi, MD[19]; Nobuyuki Miyagi, MD, PhD[20]; Toyohiro Terasaki, MD, PhD[21]; Kunihiro Kinoshita, MD, PhD[22]; Masaji Kikukawa, MD, PhD[23]; Hisakazu Kato, MD[24]; Masayuki Amano, MD[25]; Kentaro Asakura, MD[26]; Naoto Fukui, MD[27]

[1]You Home Clinic, Bunkyo-ku, Tokyo, Japan
[2]You Home Clinic Azumabashi, Sumida-ku, Tokyo, Japan
[3]You Home Clinic Azabudai, Minato-ku, Tokyo, Japan
[4]Dr. Net Nagasaki, Nagasaki-city, Nagasaki, Japan
[5]Abo Gastrointestinal Surgical Clinic, Nagasaki-city, Nagasaki, Japan
[6]Okudaira Geka, Nagasaki-city, Nagasaki, Japan
[7]Takuma Clinic, Nagasaki-city, Nagasaki, Japan
[8]Chihiro Naika Clinic, Nagasaki-city, Nagasaki, Japan
[9]Deguchi Surgery Clinic, Nagasaki-city, Nagasaki, Japan
[10]Fujii Surgical Clinic, Nagasaki-city, Nagasaki, Japan
[11]Harada Internal Medicine Clinic, Nagasaki-city, Nagasaki, Japan
[12]Nagasaki Takara Home Medical Care Clinic, Nagasaki-city, Nagasaki, Japan
[13]Nakagawa Surgical Clinic, Nagasaki-city, Nagasaki, Japan
[14]Tanigawa Clinic, Nagasaki-city, Nagasaki, Japan
[15]Ochi Clinic, Nagasaki-city, Nagasaki, Japan
[16]Nagasaki Memorial Hospital, Nagasaki-city, Nagasaki, Japan
[17]Hoshino Internal and Respiratory Medical Clinic, Nagasaki-city, Nagasaki, Japan
[18]Medical home care center, Tenri Hospital Shirakawa Branch, Tenri-city, Nara, Japan
[19]Minami Nara General Medical Center, Oyodo-town, Nara, Japan
[20]Miyagi Clinic, Tenri-city, Nara, Japan
[21]Terasaki Clinic, Nara-city, Nara, Japan
[22]Kinoshita Clinic, Sakurai-city, Nara, Japan
[23]Kikukawa Internal Medicine Clinic, Sakurai-city, Nara, Japan
[24]Kato Clinic, Uda-city, Nara, Japan
[25]Nosegawa Village National Health Insurance Clinic, Nosegawa-village, Nara, Japan
[26]Daifuku Clinic, Sakurai-city, Nara, Japan
[27]Fukui Clinic, Uda-city, Nara, Japan
